# Expert Perspectives on Clinical Laboratory Accreditation for Polygenic Risk Scores

**DOI:** 10.1101/2025.07.23.25332024

**Authors:** Courtney Wallingford, Tenielle Clinch, Lindsay F Fowles, Mary-Anne Young, Paul J. Leo, Sonia Shah, Aideen McInerney-Leo, Kirstine J Bell, Sibel Saya, Owen M. Siggs, Erika Spaeth, Emmanuelle Souzeau, Michael Inouye, Justine Marum, Stuart MacGregor, George B. Busby, Paul James, Tatiane Yanes

## Abstract

**Background:** Polygenic risk scores (PRS) are rapidly being translated into clinical practice to estimate risk to health conditions and inform health management. However, the lack of clear guidelines for laboratory accreditation of PRS restricts the ability of researchers, clinicians and regulatory bodies to evaluate test quality. Via stakeholder consultation, this study aimed to describe the unique aspects of PRS laboratory accreditation to inform test assessments by providers.

**Methods:** Eligible individuals were stakeholders involved in ordering, performing, reporting, and accrediting PRS in Australia. Semi-structured interviews explored the unique challenges and considerations of PRS testing. Interviews were transcribed verbatim, and deductive content analysis conducted.

**Results:** Participants were genetic clinicians, laboratory scientists, researchers, and industry experts with experience in PRS laboratory accreditation (n=13). Four topic areas were developed covering the entire PSR testing pipeline: i) laboratory methods, ii) PRS algorithm, iii) clinical context, and iv) reporting results. Stakeholders identified well-established laboratory methods for sample collection, genotyping and quality controls, which are applicable to PRS. Use of imputation was identified as a unique consideration for test accreditation, which impacts transparency and quality assessments of the final PRS calculation. Ancestry considerations were a major theme, impacting on all aspects of PRS pipeline. It was also widely agreed that PRS algorithms would need to be regularly identified that this process aligned with existing accreditation body allowances for test improvements. Additional considerations included the need to develop quality standards for the management of missing data, defined validation processes, and processes for collecting additional risk factors if delivering integrated risk. Finally, the importance of clear reporting of risk assessment was explored.

**Conclusions:** The findings highlight the unique considerations for PRS accreditation and can inform the development of technical standards in Australian and international diagnostic laboratories. Furthermore, a series of checklists was developed based on findings to aid providers and accreditation bodies alike assessing PRS testing.

## Background

Polygenic Risk Scores (PRS) are increasingly used clinically to estimate genetic liability to health conditions.^1^ Despite test availability, there are no established laboratory guidelines for the development and evaluation of PRS, resulting in substantial variation in the reporting of risk information.^2–4^ Variability can occur across the entire testing pipeline, including selection of risk variants (i.e. single nucleotide variants (SNVs)), algorithm development and application, missing data management and imputation, quality control steps (e.g., reference samples), and methods for addressing issues of ancestral diversity.^5,6^ For most health conditions, PRSs are likely to be delivered as part of integrated risk models that combine genetic risk information with non-genetic risk factors to deliver comprehensive risk assessments. Integrated risk models further challenge implementation due to the need for additional source data (e.g. age, lifestyle, and clinical data), which can change over time, requiring risk reassessment.^5,6^ Inconsistencies in PRS clinical reports have also been noted, including how risk information is presented, visual elements, and test limitations.^7,8^ Such variability represents a significant barrier to PRS implementation, and limits the ability of researchers, clinicians and regulatory bodies to evaluate test quality and interpret test results.

As with any clinical laboratory test, the accreditation process ensures rigor, consistency, and quality standards, thereby generating valid, reproduceable, and reliable results. Typically, accreditation is a prerequisite for a test to be eligible for public health funding (e.g. Australian Medicare services)^9^ and/or health insurance rebates. Accreditation bodies are typically country specific. In Australia, The National Association of Testing Authorities (NATA), is the leading accreditation body overseeing laboratory proficiency standards.^6^ Accreditation through NATA requires formal written applications and site visits to ensure compliance with relevant criteria, where laboratories must demonstrate proficiency standards encompassing resources, processes, and system management requirements.^10^ To date, four Australian laboratories have successfully obtained NATA accreditation for PRS tests (accreditation/site number: 21401/25782, 20432/24278, 14332/24460 and 3427/3420) for various cancers, heart disease, type 2 diabetes and glaucoma.^11^ Despite adherence to NATA criteria for human pathology testing, substantial variability remains due to the lack of PRS-specific considerations. It remains unclear how existing criteria could be adapted to capture the nuances of the PRS test and ensure consistent accreditation processes. Through consultations with key expert stakeholders, the present study aimed to identify the unique considerations for laboratory accreditation of PRS within the Australian context. Findings from this study may be used to guide future PRS laboratory accreditation guidelines and facilitate testing implementation.

## Methods

### Study design

Semi-structured interviews were conducted to capture expert stakeholders’ current practices with PRS development, experiences with laboratory accreditation, and insights into specific considerations and challenges for PRS testing regulation. Informed by the literature,^3,5,6,10,12^ a preliminary, high level, overview of the accreditation process and specific considerations for PRS was developed, which served as the foundation for constructing the interview guide (Supplementary Material 1).

### Recruitment and data collection

Eligible stakeholders were individuals working in clinical genetics, laboratory, and research roles with expertise in PRS from Australia and internationally, as well as experts in the laboratory accreditation process. An initial list of key stakeholders was developed by the research team and a snowball approach was used for additional recruitment. Eligible stakeholders were contacted via email invitation to participate and provided online consent. Interviews captured stakeholders’ experience with PRS and laboratory accreditation, including challenges related to PRS development, evaluation, interpretation and reporting (Supplementary Material 1). Interviews were audio recorded and transcribed verbatim.

### Data analysis

The qualitative analysis software NVivo (Version 14) was used to facilitate coding of interviews, and a deductive content analysis was conducted.^13^ An initial coding tree was developed to capture topics related to sample collection, genotyping, PRS algorithm development, strategies to address ancestry, use of reference samples, correcting for missing data, and clinical reporting. All transcripts were double coded by authors TC and CW to ensure rigor, and any discrepancies in coding were discussed and resolved with author TY. Initial codes were then grouped into latent themes by authors TC, CW and TY, with illustrative quotes selected. Stakeholders were invited to review findings via two rounds of consultations and contribute to the manuscript as a form of respondent validation to further enhance rigor.^14^ Representative quotes are provided to demonstrate key findings, with pseudonyms provided.

## Results

Twenty-two individuals were invited to participate in the study, and thirteen agreed to be interviewed (interview length M=47 minutes, range: 38-60 minutes). Of the nine individuals who did not participate, two were due to scheduling conflicts, one suggested an alternate expert, one was on sabbatical, and the remainder did not reply to the invitation. Expert stakeholders who participated in the study included genetic clinicians (n=5), laboratory scientists (n=3), clinical researchers (n=3), and industry experts (n=2). Of the 13 stakeholders, five had experience gaining and/or evaluating applications for PRS laboratory accreditation in Australia. Stakeholder insights were categorized into four themes pertaining to different aspects of PRS testing namely: laboratory methods, PRS algorithm, clinical context, and reporting of PRS results (Figure 1). Considerations for diverse ancestries transcended all themes, and representative quotes are presented in Table 1.

**Figure 1:**
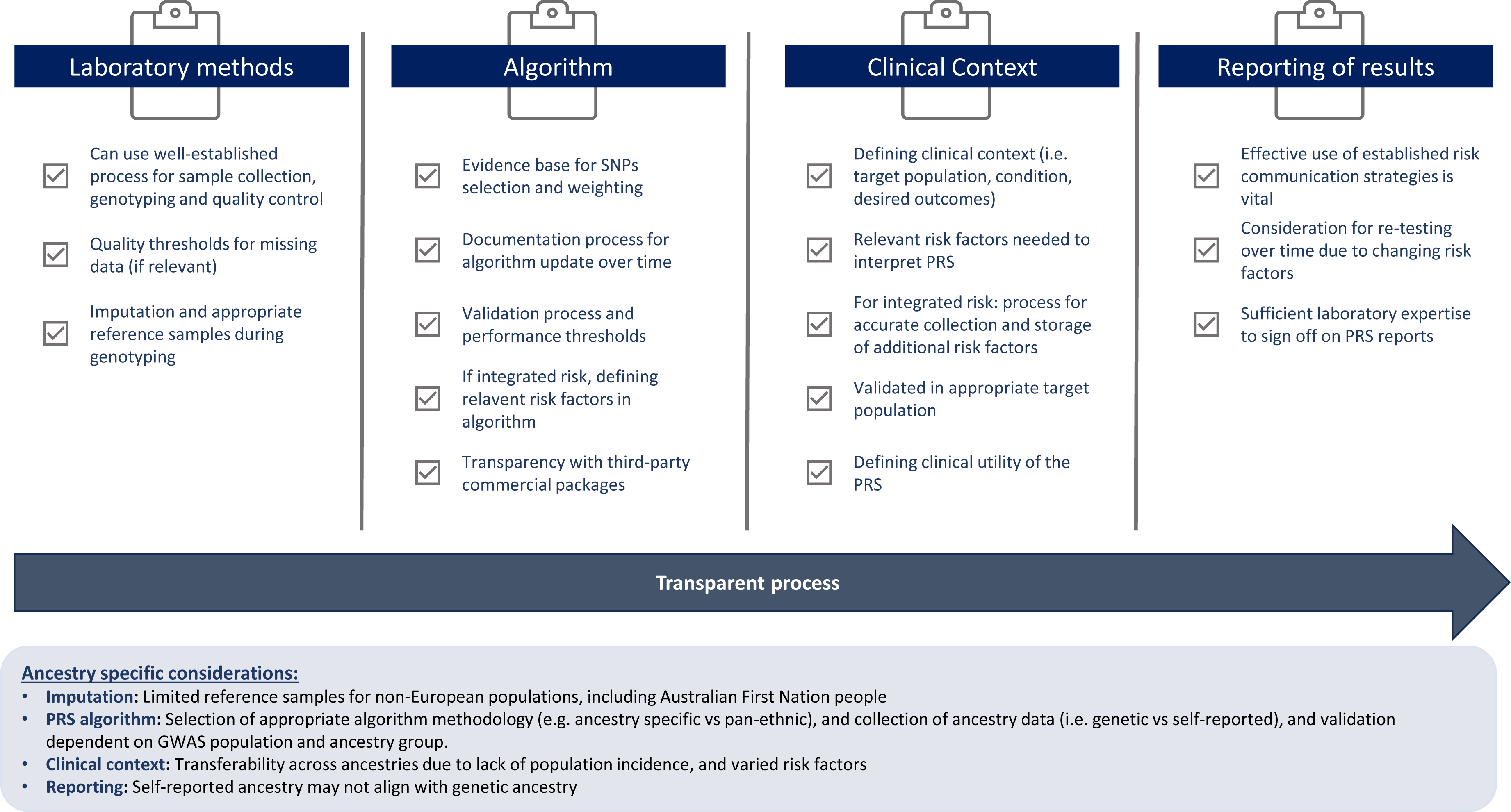
Laboratory accreditation checklist of considerations for polygenic risk scores.

**Table 1:** Themes related ancestry considerations and representative quotes.

| Considerations | Representative Quote |
| --- | --- |
| **Impact on imputation** | **Expert H:** *"It'll [ancestry] make a huge difference, and it means that imputation-based tests will be even more skewed against those underrepresented..."* |
| **Lack of population reference samples** | **Expert L:** *"And for many common ancestries, well European ancestries, it'll very, likely be fine. But if you're talking about people, a lot of Pacific nations don't have representation in a lot of the reference panels. So how to handle those samples ... what's the scope of the accreditation and what that means for equity...how to handle those individuals."* |
| **Accreditation of ancestry specific PRS** | **Expert A:** *"When we apply a test, we perform a kind of genetic ancestry assessment ... we identify the reference population that we have at our disposal that individual is most genetically similar to, so that we can then apply an ancestry PRS to that individual on the basis of that ancestry, ... So, then the complexity of regulation or accreditation around that sort of approach [ancestry specific PRS] is more difficult because you don't just have one score, you've got multiple different polygenic risk scores that you're using in different scenarios..."* |
| **Cost effectiveness of multiple PRSs** | **Expert E:** *"...how does all of this affect the cost effectiveness as well? If you run everything through one pipeline and it's very streamlined versus if you've got multiple caveats and multiple, if that then do this. If not this, then we could potentially not be able to do anything because we made it too hard..."* |
| **Defining an individual's ancestry** | **Expert H:** *"... exactly how do we define a population because there is no easy solution here. I mean humanity is not neatly divided up into different categories that correspond to clusters. We're part of a ...big continuous spectrum that it spans across the whole of humanity. And in practice what we end up having to do is to draw fairly arbitrary lines around groups of people ... there is no clarity around exactly where you draw the line..."* |
| **Lack of population specific incidence rates in Australia** | **Expert F:** *"...if a person says that they're black, we'll use incidence rates for blacks to give them their 10-year risk because for some diseases they might be at a much higher risk than white people. So, we would have to use rates that are appropriate to them...Australia doesn't have specific rates for different people."* |
| **Reporting genetically determined ancestry** | **Expert E:** *I think that's where I think you're getting into some sticky situations and if you add SNPs to genetically determine ancestry and that doesn't match what someone viewed themselves or just can you run a genetic specific risk score without returning which risk score was used?* |

### Laboratory methods

**Table 2:** Themes related to laboratory methods and representative quotes.

| Considerations | Representative Quote |
| --- | --- |
| **Algorithm development and validation** |  |
| **Selection of SNPs and weighting** | **Expert G:** *"I think you'd have to start with published data, right? I don't know many clinical laboratories who would go about creating their own [PRS]. I mean, that's a research undertaking. So then choosing which one? I think it's essentially a literature review..."* |
| **The number of SNPs included algorithm** | **Expert D:** *"...some [PRS] that get published that have 6 million SNPs. And there is no way in the world that anyone could implement a 6 million SNPs in a polygenic score without skimping on the QC and all that sort of thing because we check every one of our SNPs and make sure that they're behaving properly. So, if you get to the stupidly large PRSs, which may have slightly better performance, then I will be questioning how well they're checked."* |
| **Genotyping approach** | **Expert H:** *"...there's plenty of room for further declines in the cost of whole genome sequencing...where it becomes competitive with genotyping arrays and there's all this complexity around things like data storage, which is so much more expensive...plus processing other things more complicated..."* |
| **Updating the algorithm will require regular reporting** | **Expert C:** *"...laboratories are constantly trying to improve their assays and there's a process around this if you've been accredited you are permitted to continually improve your assay, but you have to do internal revalidation of all of those...but the moment you start going to new territory, new algorithms, new methods, you must alert them before you reintroduce it and they have the right to say, oh it's okay, or we'll do a bench review or no, no, we want to go out and do an onsite assessment."* |
| **Acceptable thresholds of accuracy** | **Expert H:** *"...what level of accuracy is sufficient, or what accuracy and benefit is sufficient to cross the line to make this an appropriate clinical test? I think it's clear that there is a substantial loss of information that happens when you take a score that was trained in one population and try to apply it to another population..."* |
| **Use of commercial packages** |  |
| **Reduce burden on laboratories** | **Expert G:** *"Ease of implementation is a really good thing if they've [commercial company] done a lot of the intellectual thinking...the ease is probably a really big pro, and if you can just upload some data and it spits something out at the end..."* |
| **Transparency** | **Expert L:** *"They're [PRS] developed by the companies, so it's sort of a bit of a black box obviously. Part of that is their intellectual property where they don't actually release the variance in weights, et cetera, that are needed to reproduce the score and recalculate it independently. So, some of that inhibits the independent assessment of the score to establish validity and utility."* |
| **Privacy considerations** | **Expert C:** *"...there are very significant accreditation considerations...Are you actually sending your patient's information to a third party? Are they accredited? If they are accredited under what system?... if you send it out, your patient relies on their accreditation. If you ingest their stuff into your system, you are acquiring it under your accreditation."* |

Overwhelmingly, stakeholders highlighted that laboratory accreditation for PRS would use the same, well-established laboratory methods for sample collection, genotyping and quality controls as applied to current monogenic genetic testing (e.g. next generation sequencing and array technology). However, stakeholders described a unique consideration for PRS quality control regarding missing data and reportable results. They emphasized the need for clearly defined thresholds and cut-offs, which considers both the total number of SNVs that could not be genotyped and the weighting of the missing SNVs, noting that the individual SNVs are not all equally important to the PRS. As noted by expert K:

> *“You can quantify how many points are missing, but you couldn’t easily quantify the influence those missing data points would have on your PRS. I mean, most have a very small influence, but some proportionately have more than others… So, you could quantify it, and you could define some sort of threshold of acceptable missing data.”*

### Considerations for imputation

Stakeholders resoundingly agreed that imputation presented unique challenges and considerations for PRS testing accreditation, with implications for transparency and quality assessments of the final PRS calculation. Stakeholders emphasized the benefits of including imputation in the genotyping process for PRS, citing that when done properly, imputation provides more accurate risk estimates, when compared to only using directly genotyped SNVs. However, the lack of representation from diverse populations in GWAS and reference samples were frequently noted as a key limitation to the imputation process (Table 1). This limitation could compromise the validity and utility of imputation for diverse ancestries, thereby potentially exacerbating health disparities. When asked about whether there was a need for Australian reference samples, stakeholders noted that they may not be necessary for the entire population but would most greatly benefit Indigenous Australians and Pasifika individuals. These populations embody significant genomic diversity relative to white-European populations and have little to no representation in currently available databases.^15,16^ However, it was equally noted that it may not be feasible to develop sufficient large databases for minority populations, and risk assessment may need to rely on novel computational approaches for recalibration.

### PRS algorithm

**Table 3:** Themes related to PRS algorithm and representative quotes.

| Considerations | Representative Quote |
| --- | --- |
| **PRS as one of many risk factors** | **Expert F:** *"...a lot of diseases have other risk factors that are really strong and should also be taken into account... you need to know at the bare minimum how old is that person...[you] could have two people with the same PRS, same family history, but if one is very young, then their risk is almost nothing. But if the second person is a bit older in their fifties or sixties, their risk could be massive."* |
| **Infrastructure for collecting clinical risk factor data** | **Expert G:** *"And then what auxiliary data do you need? Are these polygenic risk scores purely genetic or do you need some clinical data that sort of comes into those score assessments? And as soon as you need clinical data, then you need really solid processes in terms of collecting those, which I think can get laborious sometimes when you implement in a clinical setting."* |
| **Defining scope of testing** | **Expert G:** *"I think it's about defining the population that you will test... Are you testing a population with a known family history and maybe you haven't identified a rare highly penetrant variant? Or are you testing the people who already have a variant? Then you've stratifying their risks. So, I guess that all comes down to scope of testing...."* |
| **Accreditation for PRS vs integrated risk** | **Expert J:** *"...I'm sure that will be a separate accreditation process, so taking the NATA accredited genetic score result and then combining that with clinical risk factors or imaging data or whatever else, that'll be further down the line. But I think that's a really important consideration for anyone who's offering these tests."* |
| **Considerations for PRS clinical utility** | **Expert J:** *"... from the early academic work there was evidence that it was going to be clinically useful...but (sic.) for us to be able to do the trials to show that it's useful, we need an accredited test. So that's a bit of a catch twenty-two."*<br>**Expert D:** *"...we've been providing [NATA] evidence mainly from published papers that the PRS is useful, that it separates medium, higher, low risk people...It's not like it's definitely worked because I guess it's one thing that's been a little bit tricky in terms of the accreditation. It's going from a diagnostic test...to a risk stratifying test, and so it's a little bit more difficult. It's not like you can detect the variant or you can't, it's more do you get a robust score..."*<br>**Expert K:** *"even from the NATA standpoint and showing utility is not possible at a population health level because to actually show utility of a risk assessment tool would take a 50-year longitudinal study of over a million patients to be able to say that I've stratified the population now I've given them other screening options and now I've watched them until they die. And that's not a viable option. So again, the burden of proof for utility has to be lower for a risk stratification tool and that's where we're relying on solid validation data from the original design of the algorithm."* |

Stakeholders discussed many unique facets pertaining to the PRS algorithm. Considerations for PRS accreditation were detailed across the algorithm development and validation, the benefits and challenges of outsourcing the PRS calculation via commercial packages (Table 3) and adjusting for diverse ancestries (Table 1).

### Algorithm development and validation

The SNV selection process and rationale was considered as an important step in the accreditation process given its implications for the mode of sample collection, genotyping/sequencing methods, data storage and test performance, including across different ancestries. Stakeholders generally reported that they selected their PRS algorithm (including the selection and weighting of SNVs) according to the relevant literature/published GWAS data, with a preference for the largest and most recent studies/meta-analyses. The practicality of producing a large ‘extended’ PRS (i.e. including thousands or millions of variants) versus a restricted PRS (i.e. restricting inclusion to only those variants that reach genome-wide level of significance) was discussed, with many reporting this decision was dependent on costs, complexities of each approach, and target condition, noting that PRS can have a minimal impact for some conditions.^17,18^ It was also widely agreed that PRS algorithms would need to be regularly updated and refined to incorporate newly identified SNVs. However, stakeholders felt that this process aligned with existing accreditation body allowances for test improvements. Specifically, minor changes to weighting, and selection of SNVs would only require laboratories to update documentation and report back to the authority, as opposed to repeating the accreditation process (Table 3).

Independent validation of PRS algorithms was described as an essential step in the laboratory accreditation process, including reproducibility (analytical validity), and the test’s ability to accurately predict the risk or outcome (clinical validity). Stakeholders with experience with PRS accreditation described the validation stage as a “*fairly straight forward*” (Expert K) process. Analytical and clinical validation was achieved by replicating samples and repeating each step to demonstrate consistent scoring, and using case-control studies, respectively. One stakeholder emphasized that regulatory bodies: “*just care that you’re getting a reproducible result there*” (Expert D). However, stakeholders emphasized that “*acceptable*” accuracy and performance of a PRS test will depend on the relevant GWAS population, the ancestry of the individual, and the condition to which it is being applied. Therefore, agreement upon a general minimum quality and performance threshold for accreditation is not feasible, and instead dependent on the unique context for each test including purpose, setting and population) (Table 3).

### Use of commercial packages

Stakeholders commented that commercial third-party packages were increasingly available for pathology laboratories to deliver PRS and integrated risk. Many felt these packages would remove the burden of conducting statistical calculations from the laboratory, which has traditionally been beyond the scope of genetic pathologists’ expertise. However, concerns were noted about the transparency of packages, and the clinical accreditation thereof, if the algorithm could not be reviewed to allow for independent validation and quality assurance. Additionally, stakeholders mentioned concerns about the handling of patient samples/data, and liability when using data derived from a third-party commercial package (Table 3).

### Adjusting PRS for Ancestry

All stakeholders viewed ancestry as a major consideration for PRS algorithms and felt that it would be necessary for accreditation bodies to consider how this issue is addressed (Table Two broad strategies for approaching ancestry in the PRS algorithm were described: i) population-specific PRS where scores are developed based on ancestry specific data, and ii) a pan-ancestry PRS, whereby scores are derived from population data. No approach was seen as superior, with benefits and limitations noted for each. Considerations of the approach used may include:

- **Quality of ancestry-specific data:** availability of large GWAS, training data sets and reference samples.
- **Collection and definition of ancestry information:** how information is recorded (e.g. self-reported vs genetically inferred), and considerations of admixture and the spectrum upon which ancestry exists.
- **Investment in multiple test pipelines**: accreditation requirements and processes for each ancestry group and cost/resource effectiveness.
- **Ethical considerations of generating genetically inferred ancestry results:** use and privacy of information, transparency, and potential obligations and impacts of reporting back to patients.

### Clinical use of PRS: Importance of context and clinical utility

**Table 4:** Themes related to clinical practice and representative quotes.

| Consideration | Quotes |
| --- | --- |
| **Understanding the target audience** | **Expert C:** *"And so, part of the [accreditation] process, you'll be asked the question, who you provide this test to? Who is your referral market? What is their degree of familiarity with genetics, with polygenic, with statistics, show me some reports. Would they be sufficiently clear and unambiguous? Do they have enough information? Do they require supplementary information? So, you have to address those points..."* |
| **PRS is just another risk factor** | **Expert L:** *"So my view on this is that reporting should incorporate polygenic scores as just another risk factor and should be incorporated alongside existing reporting lines..."* |
| **Genetic exceptionalism and paternalism for PRS** | **Expert L:** *"...there are preexisting standards in terms of how you incorporate new risk factors that kind of transfer over directly to this sort of a setting that I would hope would be applied as well...we don't necessarily want to create a double standard for genetic risk factors, at least when it comes to risk prediction because we kind of know how to incorporate new risk factors. It happens all the time..., but having double standards for polygenic score, sort of genetic exceptionalism and that's not something that you want..."* |
| **Clear reporting is essential** | **Expert C:** *"Researchers are very good at coming out with excellent reports that are utterly unintelligible to the average clinician...depending on the market, it may be one-page front summary. Your result is, here's a nice picture, you can download the other 16 pages here..."*<br>**Expert B:** *... having different types of visual aids ... your bell curve and absolute risk, that was either represented as a pie or as your icon array where you have all the little individuals represented out of a hundred who actually get the disease... People prefer absolute risk to relative risk... and we can still include relative risk, we can still say, look, your risk is two or three times higher in the general population."* |
| **Dynamic reports not feasible** | **Expert C:** *"Once you issue it, you can't have a dynamic report. It has to be locked in time as it was at the moment you authorized and issued it. If you're going to change it or update it, you have to issue supplementary reports. You can't have the report changing in time."* |
| **Education and training for clinicians would be beneficial** | **Expert G:** *"The whole part of clinical governance and oversight is that you have training and competencies, and you have qualifications that go behind people who analyze and sign things out...it's actually part of NATA assessment and accreditation is that you need to make sure that you have competent scientists. So, I don't think there's a specific qualification, but in general, obviously quite similar to any genomic test, you've got to have the relevant experience... I think it comes down to training, competency assessments and whatnot, which is pretty standard for clinical diagnostic laboratories."* |

Stakeholders’ discussions about PRS accreditation and clinical use centered around the importance of contextualizing the information and ensuring clinical utility (Table 4). Unanimously, stakeholders believed that PRS should be viewed as one of many risk factors, which can be combined with monogenic testing and other non-genetic risk factors (i.e. integrated risk). Furthermore, stakeholders reported that for most conditions, it was neither clinically useful, nor responsible to interpret PRS in isolation. For example, one participant stated that age and sex, “*at the bare minimum*” must be considered to provide absolute risk for a specific timeframe. Therefore, regulatory bodies will need to consider how the PRS will be applied, and if calculating integrated risk, ensure the necessary processes are in place to accurately capture the required clinical data (e.g. family history, lifestyle factor, imaging data and/or biochemical markers). While integrated risk provides more comprehensive risk assessment, limitations of this information were noted, such as the transferability across ancestral groups due to lack of population-specific incidence rates and varied risk factors (Table 1). Thus, stakeholders highlighted the importance of defining the scope and purpose of the test, including identifying the target population, desired outcomes (e.g. personalized screening regimes for early detection or lifestyle changes for prevention) and test limitations. Of note, many stakeholders felt that accreditation of PRS would be a separate process to that of an integrated risk, but some stakeholders detailed that they had, in fact, obtained laboratory accreditation for an integrated risk score.

The capacity to demonstrate clinical utility was identified as a necessary part of the Australian NATA accreditation process. However, stakeholders highlighted that benchmarks for clinical utility will vary based on conditions and settings for which PRS is being used. The need to consider PRS as population screening tool was also noted, which represents a shift from the traditional diagnostic genomic testing. Stakeholders recognized that clinical risk thresholds and corresponding advice/interventions did not currently exist for many conditions, further complicating the evaluation of clinical utility. Lastly, evidence of clinical utility for population-wide risk stratification tests was described as a complex process that could take decades. Despite these challenges, stakeholders were not concerned by the limited evidence for real world clinical utility given availability of strong modelling data for specific conditions. Many believed this information could be gathered alongside testing accreditation and implementation.

### Reporting and communicating PRS results

**Table 5:** Themes related to reporting of results and representative quotes.

Effective risk communication to both clinicians and patients was identified as an essential consideration for the accreditation process. Laboratory scientists experienced in clinical accreditation noted that authorities require details about the nature, quantity, and mode of result communication, and the characteristics of the target audience. Nonetheless, stakeholders emphasized that PRS is “*just another risk factor*” and that it should be communicated and evaluated using the same established strategies for effective risk communication. Stakeholders described a wide range of possible approaches for risk communication that included presenting information in different formats (i.e. visual, numeric, or categorical) and using different numeric risks such as absolute and relative risk, with a noted preference for the former. Despite this belief, some stakeholders cited concerns and experiences with paternalism and double standards for PRS (and genetics more broadly) as compared to traditional clinical risk assessment tools. Additionally, some stakeholders suggested that integrated risks be communicated using dynamic reports that can be updated to reflect changes in non-genetic risk factors over time. However, others noted that this approach would not be possible as accreditation bodies prohibit the alteration of reports once issued and signed. Thus, clinical providers need to request new testing and provide updated non-genetic risk information (Table 5). Similarly, reports may need to be revised due to changing PRS algorithms, and it was noted that processes would need to be in place to accommodate this revision. Stakeholders suggested that this could include automatic reissuing of reports or relying on clinicians to request updated recalculations.

There was consensus that clinical reports should include details of the methods used for generating the PRS (i.e. the panel, imputation software/version, and algorithm used), risk factors included (if integrated risk), and test limitations. Stakeholders felt that accreditation bodies should avoid mandating the reporting of specific numbers or values, as it may not be practical or useful to patients and/or clinicians in specific contexts (Table 5). Some stakeholders also raised concerns about the ethical challenges of reporting information such as genetically determined ancestry. Specifically, they detailed that this information may need to be disclosed for transparency and reproducibility, but ethically, the information may be confronting patients if it does not concur with their identified ancestry (Table 1).

Stakeholders agreed that the process of generating PRS reports, as well as determining who would be qualified to sign off on results in the laboratory would remain unchanged from current genomic testing processes. However, it was acknowledged that part of the current accreditation process for human pathology testing mandates that the laboratory requires the appropriate expertise to sign off on the report. Thus, stakeholders collectively felt that additional training may be required to support genetics pathologists and clinicians working in laboratories seeking clinical accreditation for PRS testing (Table 5).

## Discussion

This study describes the unique nuances for PRS laboratory accreditation as identified by expert Australian stakeholders (Figure 1). Many of the challenges and considerations identified are not unique to PRS, but reflect broader systemic issues, such as the underrepresentation of diverse ancestries and the consequent poorer outcomes across healthcare settings.^19^ Another potential challenge is the evolving PRS algorithms and variable risk estimates that can result in shifting risk classification. Similar issues occur in diagnostic genomic testing due to changes in variant pathogenicity classification and test re-analysis, which are subject to laboratory specific protocols.^20^ Nevertheless, several unique PRS considerations were noted including issues related to imputation and management of missing data, complexities of integrated risk calculations, and workforce training needs.

A need for clear and transparent processes across the PRS pipeline consistently identified as an area of priority for laboratory accreditation. Given the broad potential uses of PRS-based risk assessment, it is critical that laboratories seeking clinical accreditation provide clear information on the clinical context of the intended test, such as target population, condition, and desired purpose. Such transparency is also essential to ensure healthcare providers and patients alike have the necessary information to evaluate and compare testing. Furthermore, a key consideration for PRS testing is the use of integrated risk models and inclusion of non-genetic risk factors that can substantially alter final risk assessments.^3,6^ For example, breast density is one of the strongest independent predictors for breast cancer, and thus, frequently included in integrated risk models for this condition.^21^ However, historically breast density has not been routinely reported to women undergoing population breast screening in Australia. Consequently misconceptions about breast density have been reported among the public that can potentially result in inaccurate integrated risk assessment if reliant on patient-self report data.^22,23^ Thus, laboratories providing integrated risk will require clear processes and regulations for the accurate collection, management, and integration of additional risk factors. ^3,6^ Finally, consideration is needed regarding whether genetic pathologists require additional training to ensure capacity to sign off PRS test reports given that this test is not currently included in their competency criteria.^24^

In line with international literature,^25,26^ ancestry considerations were a major theme of this study that impacts all aspects of the PRS and integrated risk pipeline (Figure 1; Table 1). Ancestry considerations stemmed from limited diverse data across genomics databases and epidemiological studies that are used to inform integrated risk calculation (i.e. population incidence data and population-specific risk factors).^19^ While there is extensive work within the genetics community to improve the current lack of ancestral diversity of genomic databases, limited progress has been made to date.^27^ Currently, there are no ‘best practice’ methods established to address the ancestry related issues, with each individual laboratory making decisions based on their unique context and preferred approach. Thus, accreditation bodies should be aware of the different approaches and limitations to ensure appropriate regulation of PRS testing. In instances where genetic ancestry inference is used, transparency must be maintained for interpretation and validation purposes.^28^ However, previous studies have highlighted the potential for inferred genetic ancestry to result in both benefits and harms (i.e. more personalized risk prediction versus risk of mismatched self-identified and inferred genetic ancestry), and advised that stakeholders should be afforded choice about receiving this information.^28,29^ Therefore, a comprehensive informed consent process is warranted, outlining the information that will be returned in regards to the PRS generation process, including uncertainties, and limitations.

Communication of PRS results represents a shift from the traditional reporting of Mendelian genetic tests to an algorithm-based test that requires interpretation of complex risk information and consideration of epidemiological risk modeling. Accurate and understandable reporting is thus vital to ensure healthcare providers and patients can interpret and make informed decisions from test results. The essential nature of clear reports is further emphasized given the lack of knowledge and confidence regarding PRS frequently reported by healthcare providers.^30–32^ Determining best practices in PRS and integrated risk reporting is an area of active research.^7,8,32,33^ However, given the broad potential applications for PRS testing, it is unlikely that a single best practice method will be universally developed. Nevertheless, key concepts of complex risk communication can be applied including, outlining risk in multiple formats, using absolute risk over relative risk, contextualizing risk estimates, and limiting the amount of numeric information conveyed.^8,34–36^ Clinical workforce education and support to uptake appropriate clinical risk management strategies are also needed.^37^

Further research and consultation are needed to develop standardized pathways and processes for PRS laboratory accreditation in Australia. Based on the findings from this study, we propose the development of an Expert Advisory Group to refine guidelines for PRS accreditation process, while ensuring transparency and national standards. This process should encompass engagement with relevant accreditation and regulatory bodies, and other key stakeholders (i.e. healthcare providers, laboratory services, legal experts and consumers) to establish standards that not only align with current accreditation criteria but also address the specific nuances and unique considerations necessary for PRS clinical laboratory accreditation. Furthermore, developing a centralized resources repository is recommended to support accreditation bodies and healthcare providers alike to review and interpret information derived from this new test.

Strengths of this study included comprehensive stakeholder engagement through an expert committee, which informed the development of a detailed conceptual overview of the PRS testing process and the semi-structured interview guide. Findings were also reported back to stakeholders for clarification and additional input on the PRS accreditation process.

Limitations included the lack of broader community perspectives, such as those from patients or policymakers, and the use of a snowball recruitment method, which may have introduced selection bias, resulting in a more homogeneous cohort with similar viewpoints. Nevertheless, this study comprehensively explored the unique considerations for PRS testing and laboratory accreditation in Australia. Key areas highlighted include quality control, imputation, algorithm development and validation, appropriate context, and standardized reporting and communication. These findings can inform future research, guideline development for PRS accreditation, and educational modules for laboratory services, and aid clinicians and patients ordering and interpreting results.

## Data Availability

The datasets generated and/or analyzed during this study include semi-structured interview transcripts that contain sensitive participant information and cannot be shared publicly to protect participant privacy. However, the coding trees derived from these interviews are available from the corresponding author upon reasonable request.

## Declarations

### Ethics approval and consent to participate

This study was approved by the University of Queensland Human Research Ethics Committee (2023/HE000986). All stakeholders provided written informed consent prior to inclusion in the study and consented to the publication of anonymized data in scientific journals.

### Authors’ contributions

Funding acquisition: PJ and TY. Conceptualization: MAY, PJL, SS, AML, PJ, and TY. Data collection: TC, LFF, AML, and TY. Initial analysis: CW, TC, MAY, SS, PJ, and TY. Writing original draft: CW and TY. Draft review and result synthesis: all authors. All authors read and approved of the final manuscript.

## Acknowledgements

We thank all individuals who participated in the study and provided input into PRS test accreditation. We also gratefully thank Amali Disanayaka from Australia Genomics for her initial support in the project, including assistance in scheduling interview and study administration.

